# Nurses’ Support of Treatment Decision-Making for Patients with Cancer During the COVID-19 Pandemic in Japan

**DOI:** 10.1101/2024.06.01.24308318

**Authors:** Shiori Yoshida, Konosuke Sasaki, Fumiko Sato

## Abstract

During the COVID-19 pandemic, cancer patients became reluctant to come to the hospital, receive cancer treatment, and were willing to interrupt or postpone treatment due to concerns about infection. The purpose of this study was to discuss effective treatment strategy decision making support for cancer patients by nurses recognized during the COVID-19 pandemic. The study population comprised nurse of decision-making support at cancer care during COVID-19 from May to July 2021 at 49 the cancer care center hospitals were established in each prefecture, who had engaged 50% of their nursing care provided to patients with cancer.

Data were collected on treatment strategy decision-making support using an online cross-sectional survey. Factors that influenced patients’ decision-making were identified using multivariable logistic regression analysis. A total of 182 (25.0%) were nurses engaged in decision making were enrolled in this study. Factors that influenced patients’ decision to receive treatment to their satisfaction were their resignation or compromise in accepting the decision due to the pandemic (odds ratio [OR] 0.44 (95% CI [0.22, 0.87]), videoconference follow-up (OR 0.17, 95% Cl [0.04, 0.66]), and infection prevention information provision (OR 3.82, 95% Cl [1.54,9.46]). Factors influencing patients’ decision to give up and accept the doctor’s recommendation even though they were not convinced included fear of disease progression (OR 2.51, 95% Cl [1.21, 5.22]), anger at not receiving the treatment they desired (OR 2.48, 95% Cl [1.17, 5.27]), and compromise with the pandemic situation (OR 3.15, 95% Cl [1.53, 6.50]). The factor that influenced patients’ continued treatment even though they were not convinced included the nurse listened to the patients’ thoughts (OR 0.07, 95% Cl [0.01, 0.51]). Treatment decision support of patients with cancer during COVID-19 included lifestyle guidance to prevent the spread of infection and listening to their concerns to help them find meaning in their choices.

## Introduction

COVID-19 was first identified in China in December 2019 and spread rapidly worldwide. The World Health Organization ^1)^ characterized it a global pandemic on March 11, 2020, declaring the end of COVID-19 as an international public health emergency in 2023. As of last reported on September 1, 2023, more than 770 million cases and 6.9 million deaths were reported worldwide^2)^. Patients with cancer have a 3.5-fold higher risk of contracting a serious COVID-19 infection ^3)^ and a 5.6% risk of fatality ^4)^. Nevertheless, they must make treatment choices even during a life-threatening crisis, such as the COVID-19 pandemic. Medical treatment strategies include adjuvant chemotherapy and the postponement of surgery for low-risk patients with cancer^3)5)^.

Physical distancing (sometimes called social distancing) became the norm during the COVID-19 pandemic^6)^. Consequently, patients with cancer refrained from visiting the hospital due to fear of infection and were reluctant to receive cancer treatment, with some even considering suspending or postponing their treatment ^7)^. In Japan, cancer screening uptake declined by 10%–30% during the pandemic period^8)^, and cancer diagnosis registrations decreased by 4.6%^9)^. This was due to delays in screening and receiving medical care as a result of new COVID-19 infections and not due to a decrease in the number of cancer cases^10)^. Similarly, cancer center hospitals in Latin America reported a 60% decrease in the number of surgeries, a 27.5% decrease in patients receiving intravenous chemotherapy, and a 30.9% increase in new patients receiving oral chemotherapy compared to the pre-COVID-19 period ^11)^.

The COVID-19 pandemic caused patients with cancer to have increased anxiety regarding what they can do to protect themselves and how their treatment would be affected^12)^. Depression—which reduces survival in patients with cancer^13)^—was reported to be 8%–24% among this cohort of patients before the pandemic^14)^ compared to 40.7% during the pandemic^15)^. Patients’ psychological distress can create conflicts in their treatment strategy selection^16)^. Discussions between physicians and patients regarding receiving cancer treatment during the COVID-19 pandemic were potentially psychologically distressing because of the added life-threatening risk of infection, differing from discussions regarding the usual treatment. However, it was unknown how patients with cancer would react and what kind of treatment plan they would choose in the event of a pandemic. Furthermore, the actual state of nursing care to support their decision-making was unclear.

Using an online survey, we investigated the actual decision-making support provided by nurses to patients with cancer during the COVID-19 pandemic, which often required considering changes in treatment plans. This study provides basic data on what type of nursing care can support patient decision-making during any future pandemic. Pandemics caused by infectious diseases are classified as a disaster nursing event. In the future, our findings may guide nurses in providing decision support to patients with cancer during global health crises.

## Materials and Methods

### Study design, setting, and participants

This was an online cross-sectional study was conducted throughout Japan as COVID-19 infections were identified nationwide. Participants were nurses working in the wards or outpatient clinics of all cancer treatment base hospitals in Japan from May to July 2021, with 50% of their nursing care provided to patients with cancer. The cancer nursing engagement ratio was used as a reference for the amount of work required in the application for Certified Nurse Specialist in Cancer Nursing. In Japan, cancer care center hospitals were established in each prefecture to provide high-quality cancer care. These hospitals provide cancer care even in situations where infectious diseases are prevalent^8)^. Nurses at these hospitals were included in the study because they were expected to provide decision-making support for cancer treatment during the COVID-19 pandemic.

A survey request form and a survey instruction sheet with a QR code that could be used to download the survey form were sent to the heads of nursing managers at 49 Japanese cancer care center hospitals. Nursing managers distributed the survey instruction sheet to nurses who were willing to participate in the survey. Informed consent was provided in the survey instructions and it was explained that those who responded to the online survey were considered to have consented to the survey.

## Measures

### Socio-demographic characteristics

Socio-demographic variables, including nurse’s age, year of cancer nursing experience for nurse, emergency declaration area, situations in which decision-making support for treatment strategies was provided, department and barriers to overall cancer nursing practice caused by COVID-19.

### The decision support for COVID-19-related treatment strategies in cancer patients Questionnaire

The decision support for COVID-19-related treatment strategies in cancer patients Questionnaire items were extracted from interviews with two certified nurse specialists in cancer nursing affiliated with a base hospital for cancer care. The questionnaire covered various aspects, including determined treatment strategy(3 items), the characteristics of the patient who needed support for treatment decision as recognized by the nurse (5items), the nursing care provided to patients who needed decision-making support (13items), the patient’s response to the nursing practice as recognized by the nurses (4 items). Participants selected all that applied item.

### Statistical analysis

The data were analyzed using the IBM® SPSS® Statistics 27.0 statistical software package. The independent variables were factors related to patients’ treatment decisions, namely, “characteristics of patients needed support for treatment decisions as recognized by the nurses” and “nursing care provided to patients who needed decision-making support.” The dependent variable was “the patient’s response to the nursing practice as recognized by the nurses.”

The relationship between the factors related to characteristics of patients needing support for treatment decisions as perceived by nurses and response to the nursing practice as recognized by the nurses and the patients’ treatment decisions was analyzed by a chi-square test. A multivariable logistic regression analysis was performed using the chi-square test results for each item, with items with a *p* value less than 10% as the independent variables and “the patients’ response to nursing practice as perceived by nurses” as the dependent variable.

### Ethical consideration

This study was reviewed and approved by the Ethics Committee Tohoku University Graduate School of Medicine (Approval No. 2021-1-215). Informed consent was considered to be provided upon the completion of the online survey and to extend to the registration of the results. Responses to the survey were handled anonymously.

## Results

The survey was sent to 49 cancer treatment center hospitals in Japan, and a total of 182 (25.0%) were nurses engaged in decision making were enrolled in this study.

### Characteristics of the study participants

Table 1 shows participant characteristics. The mean number of years of nurse’s cancer nursing experience was 13.3 (*SD* = 8.52) years. Nurses provided support in the decision-making process for 61 (33.5%) outpatients and 133 (73.1%) inpatients. The most common department in which nurses worked was respiratory medicine (54; 29.7%), followed by gastroenterological surgery (41; 22.5%) and gastrointestinal medicine (40; 22.0%). The most common barrier to overall cancer nursing practice caused by COVID-19 was “lack of nursing care support for family caregivers due to restricted visitation during hospitalization” in the case of 147 (80.8%) patients. This was followed by “more unusual tasks such as infection prevention measures and zoning for COVID-19” in 135 (72.2%) and “lack of psychological support for patients” in 109 (59.9%) patients.

**Table 1.**

| *Participant Characteristics* |  | *n* | *%* | *M* | *SD* |
| --- | --- | --- | --- | --- | --- |
| Age |  |  |  | 39.7 | 10.04 |
| Nurse's cancer nursing experience (years) |  |  |  | 13.3 | 8.52 |
| Emergency declaration area |  |  |  |  |  |
|  | Emergency | 12 | 6.6 |  |  |
|  | Prevention of spread | 53 | 29.1 |  |  |
|  | Not applicable | 117 | 64.3 |  |  |
| Situations in which decision-making support for treatment strategies |  |  |  |  |  |
|  | Outpatient | 61 | 33.5 |  |  |
|  | Inpatient | 133 | 73.1 |  |  |
| Department (multiple responses) |  |  |  |  |  |
|  | Respiratory medicine | 54 | 29.7 |  |  |
|  | Gastrointestinal surgery | 41 | 22.5 |  |  |
|  | Gastrointestinal medicine | 40 | 22 |  |  |
|  | Otorhinolaryngology | 34 | 18.7 |  |  |
|  | Hematology | 34 | 18.7 |  |  |
|  | Medical oncology | 29 | 15.9 |  |  |
|  | Gynecology | 26 | 14.3 |  |  |
|  | Respiratory surgery | 24 | 13.2 |  |  |
|  | Urology | 20 | 11 |  |  |
|  | Breast | 17 | 9.3 |  |  |
|  | Radiology | 6 | 3.3 |  |  |
|  | Pediatrics | 5 | 2.7 |  |  |
|  | Other | 16 | 8.8 |  |  |
| Barriers to overall cancer nursing practice caused by COVID-19 |  |  |  |  |  |
|  | Lack of nursing care support for family caregivers due to restricted visitation during hospitalization | 147 | 80.8 |  |  |
|  | More unusual tasks such as infection prevention measures and zoning for COVID-19 | 135 | 74.2 |  |  |
|  | Lack of psychological support for patients | 109 | 59.9 |  |  |
|  | Lack of nursing care support for family caregivers due to restrictions on accompanying patients during outpatient visits | 79 | 43.4 |  |  |
|  | Limitation of nursing intervention time | 75 | 41.2 |  |  |
|  | Lack of social support for patients | 74 | 40.7 |  |  |
|  | Lack of spiritual support for patients | 56 | 30.8 |  |  |
|  | Lack of symptom management support for patients | 43 | 23.6 |  |  |

### The decision support for COVID-19-related treatment strategies in patients with cancer

Table 2 shows the decision support for COVID-19-related treatment strategies in patients with cancer. The determined treatment strategy was postponement of treatment (131; 72.0%), followed by change of treatment (71; 39.0%) and discontinuation of treatment (41; 22.5%). The most common characteristics of patients who needed support for treatment decision as recognized by the nurses were “fear of disease progression” (93; 51.1%), followed by “anxiety about the future” (89; 48.9%) and “resignation or compromise in accepting the decision due to the pandemic” (82; 45.1%). Nursing care provided to patients who needed decision-making support included 169 (92.9%) nurses who listened to the patient’s thoughts, 87 (47.8%) who listened to the patient’s intentions and thoughts again after treatment decision, and 77 (42.3%) who coordinated opportunities for additional explanations from the doctor. The response of the patients to nursing practice as recognized by the nurses included 128 (70.3%) patients who were satisfied and received the treatment they had chosen, and four (2.2%) who continued the treatment because they were not convinced.

**Table 2.**

| Table 2 | N = 182 |  |
| --- | --- | --- |
| Decision Support for COVID-19-Related Treatment Strategies in Patients with Cancer |  |  |
|  | n | % |
| Determined treatment strategy (multiple responses) |  |  |
| Postponement of treatment | 131 | 72.0 |
| Change of treatment | 71 | 39.0 |
| Discontinuation of treatment | 41 | 22.5 |
| Characteristics of patients needing support for treatment decisions as perceived by the nurse (multiple responses) |  |  |
| Fear of disease progression | 93 | 51.1 |
| Anxiety about the future | 89 | 48.9 |
| Resignation or compromise in accepting the decision due to the pandemic | 82 | 45.1 |
| Anger at not receiving the desired treatment | 54 | 29.7 |
| Anticipatory anxiety about the COVID-19-induced death of a famous person coinciding with their own death | 33 | 18.1 |
| Other | 12 | 6.6 |
| Nursing care provided to patients who needed decision-making support (multiple responses) |  |  |
| The nurse listened to the patient's thoughts | 169 | 92.9 |
| The nurse listened to the patient's intentions and thoughts again after the treatment decision | 87 | 47.8 |
| The nurse coordinated opportunities for additional explanations from the doctor | 77 | 42.3 |
| The nurse explained the physical and mental impact of the chosen treatment strategy to patients | 58 | 31.9 |
| The nurse provided information about infection prevention while living with COVID-19 | 52 | 28.6 |
| The nurse assured the patient that they could take immediate action with the doctor's permission if symptoms worsened | 35 | 19.2 |
| The nurse confirmed the reason for the selection and corrected the patient if their perception was incorrect | 32 | 17.6 |
| The nurse followed up with the patient's symptoms and changes in decision-making on treatment strategies via telephone | 27 | 14.8 |
| The nurse followed up via telephone with the patient's family caregivers | 20 | 11.0 |
| The nurse followed up via videoconference with the patient's family caregivers | 12 | 6.6 |
| The nurse followed up via videoconference concerning changes in the patient's symptoms and decision-making perceptions | 5 | 2.7 |
| The nurse provided encouragement | 5 | 2.7 |
| The nurse provided interventions to help patients gain the strength to overcome the crisis | 4 | 2.2 |
| Other | 2 | 1.1 |
| The patient's response to nursing practice as recognized by nurses (multiple responses) |  |  |
| The patient was satisfied and received the chosen treatment | 128 | 70.3 |
| The patient was not convinced, but gave up and accepted the treatment recommended by the doctor | 61 | 33.5 |
| The patient was not convinced, but gave up and decided to discontinue treatment | 12 | 6.6 |
| The patient was not convinced and continued with the previous treatment | 4 | 2.2 |
| Other | 4 | 2.2 |

### Univariate analysis of decision support for COVID-19-related treatment strategies in patients with cancer

Table 3 shows the relationship in univariate analysis between the factors related to “the characteristics of patients needed support for treatment decisions as recognized by nurses” and “the nursing care provided to patients who needed decision-making support” as independent variables, and the relationship with “the patient’s attitude to the nursing practice as perceived by nurses” as the dependent variable. The number of nurses who recognized that a patient had “resignation or compromise in accepting the decision due to the pandemic” increased significantly, indicating that patients were satisfied with their treatment (*p* = 0.04). The number of nurses who “provided information about infection prevention while living with COVID-19” increased significantly, indicating that patients were satisfied with their treatment (*p* = 0.03). However, the number of nurses who “followed up via videoconference with the patient’s family caregivers” decreased, indicating that patients were satisfied with their treatment (*p* = 0.05).

**Table 3.**
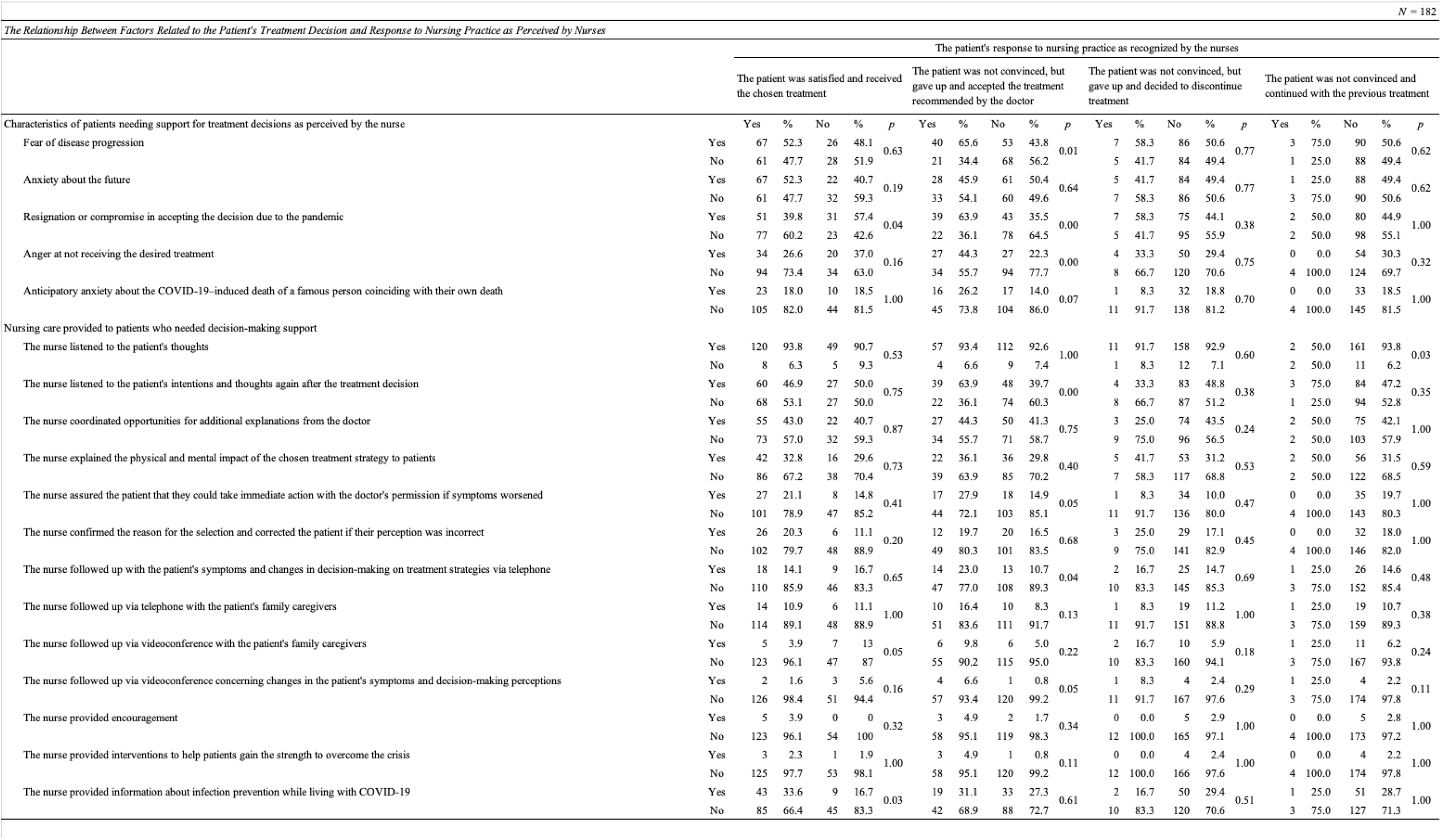

The number of nurses who recognized that a patient had “fear of disease progression,” “resignation or compromise in accepting the decision due to the pandemic,” “anger at not receiving the desired treatment,” and “anticipatory anxiety about the COVID-19–induced death of a famous person coinciding with their own death,” increased significantly, thus indicating that patients was not convinced, but gave up and accepted the treatment recommended by the doctor (*p* < 0.10 for each). The number of nurses who practiced “listened to the patient’s thoughts” decreased significantly, leading to the perception that the patient was not convinced, and thus they continued with the previous treatment (*p* = 0.03). Conversely, the number of nurses who “listened to the patient’s intentions and thoughts again after the treatment decision,” “assured the patient that they could take immediate action with the doctor’s permission if symptoms worsened,” “followed up with the patient’s symptoms and changes in decision-making on treatment strategies via telephone,” and “followed up via videoconference concerning changes in the patient’s symptoms and decision-making perceptions,” increased significantly, indicating that the patient was not convinced, but gave up and accepted the treatment recommended by the doctor (*p* < 0.10 for each).

### Multivariate analysis of LCS behaviors in high-risk individuals

Table 4 shows results of the multivariate analysis of the factors influencing patients’ treatment decisions as recognized by nurses. Factors that influenced the patients’ decision to receive treatment to their satisfaction were “resignation or compromise in accepting the decision due to the pandemic” with an odds ratio (OR) of 0.44 (95% Cl [0.22, 0.87]), “the nurse followed up via videoconference concerning changes in the patient’s symptoms and decision-making perceptions” with an OR of 0.17 (95% Cl [0.04, 0.66]) and “the nurse provided information about infection prevention while living with COVID-19” with an OR of 3.82 (95% Cl [1.54, 9.46]). These results indicate that videophone follow-up is not necessary and that the provision of infection prevention information by nurses and patient’s resignation or compromise in accepting the decision due to the pandemic influences patients’ decisions to receive satisfactory treatment.

**Table 4.**

|  |  | B | SE | OR | 95% CI | p value |
| --- | --- | --- | --- | --- | --- | --- |
| The patient was satisfied and received the treatment they had chosen | Resignation or compromise in accepting the decision due to the pandemic | -0.818 | 0.346 | 0.44 | 0.22-0.87 | 0.018 |
|  | The nurse followed up via videoconference concerning changes in the patient's symptoms and decision-making perceptions | -1.765 | 0.689 | 0.17 | 0.04-0.66 | 0.010 |
|  | The nurse provided information about infection prevention while living with COVID-19 | 1.339 | 0.463 | 3.82 | 1.54-9.46 | 0.004 |
| The patient was not convinced, but gave up and accepted the treatment recommended by the doctor | Anticipatory anxiety about the COVID-19-induced death of a famous person coinciding with their own death | 0.481 | 0.470 | 1.62 | 0.64-4.06 | 0.306 |
|  | Fear of disease progression | 0.919 | 0.374 | 2.51 | 1.21-5.22 | 0.014 |
|  | Anger at not receiving the desired treatment | 0.908 | 0.385 | 2.48 | 1.17-5.27 | 0.018 |
|  | Resignation or compromise in accepting the decision due to the pandemic | 1.147 | 0.370 | 3.15 | 1.53-6.50 | 0.002 |
|  | The nurse listened to the patient's intentions and thoughts again after the treatment decision | 0.614 | 0.370 | 1.85 | 0.89-3.81 | 0.097 |
|  | The nurse followed up with the patient's symptoms and changes in decision-making on treatment strategies via telephone | 0.082 | 0.548 | 1.09 | 0.37-3.18 | 0.881 |
|  | The nurse followed up via videoconference concerning changes in the patient's symptoms and decision-making perceptions | 1.755 | 1.306 | 5.78 | 0.45-74.75 | 0.179 |
|  | The nurse assured the patient that they could take immediate action with the doctor's permission if symptoms worsened | -0.012 | 0.476 | 0.99 | 0.39-2.51 | 0.979 |
| The patient was not convinced and continued with the previous treatment | The nurse listened to the patient's thoughts | -2.72 | 1.047 | 0.07 | 0.01-0.51 | 0.009 |

Factors influencing patients’ decision to give up and accept the doctor’s recommendation even though they were not convinced included “fear of disease progression” (OR 2.51, 95% Cl [1.21, 5.22]), “anger at not receiving the desired treatment” (OR 2.48, 95% Cl [1.17,5.27]), and “resignation or compromise in accepting the decision due to the pandemic” (OR 3.15, 95% Cl [1.53, 6.50]). These results indicate that patients were reluctant to accept recommended treatments due to fear of disease progression, anger at not receiving the treatment and their resignation or compromise in accepting the decision due to the pandemic.

The factor that influenced patients’ continued treatment—even though they were not convinced—included “the nurse listened to the patient’s thoughts” (OR 0.07, 95% Cl [0.01, 0.51), indicating that nurses’ failure to listen to patients’ thoughts is a contributing factor to patients continuing treatment even though they are not satisfied with it. “The patient was not convinced but gave up and decided to discontinue treatment” had no analysis results as no items fell below the 10% significance level.

## Discussion

This study investigated the experiences of nurses to examine effective nursing support for decision-making regarding treatment strategies for patients with cancer during the COVID-19 pandemic. During the study period, Japan was experiencing the fourth and fifth waves of the COVID-19 outbreak. This period was characterized by the emergence of new variants, such as the alpha and delta strains, which caused a critical medical situation^17)^, and both patients and medical personnel being fearful of the unknown infectious disease. Moreover, vaccine uptake for COVID-19 was only approximately 30% in Japan^18)^. During the COVID-19 pandemic, 72% of the patients in our study had to make decisions regarding postponement of their treatment, 39% of the patients chose to change treatment, 22.5% chose to discontinue treatment, 51.1% were worried about disease progression, and 48.9% were worried about an uncertain future.

### The effective ways to support of treatment decision-making for patients with cancer during COVID-19 pandemic

The findings of this study suggest that the effective ways to provide care for continuing cancer treatment included providing patients information on preventing COVID-19 infection and patients’ attitude of resignation or compromise in accepting the decision due to the pandemic. Additionally, patients’ acceptance of doctor-recommended treatment—even if they were not satisfied with it—was influenced by patient attitudes such as anger, fear of disease progression, and their resignation or compromise in accepting the decision due to the pandemic.

First, during this pandemic, patient education on living with COVID-19 infection prevention helped convince patients to accept the treatment. Patients with cancer undergoing chemotherapy or surgical treatment are at an increased risk of contracting COVID-19^3)^, which in turn increases the risk of morbidity and mortality ^19)^. During the decision-making process, patients may change their treatment strategy based on the impact of the change in treatment on the deterioration of their condition and the risk of contracting COVID-19 and dying. One of the reasons for refusal of treatment due to COVID-19 was the risk of infection ^20)^. Patients with cancer with a high fear of infection tend to delay treatment ^21)^. Thorough infection control measures in the hospital and lifestyle guidance for patients to prevent the spread of infection are part of the basic care that patients need when trying to decide on a treatment plan with which they are satisfied. Another factor influencing patient satisfaction with decision-making was the lack of a need for follow-up by nurses via video calls. Telemedicine was rapidly introduced during the pandemic, but one report found that video calls were inefficient when used to explain uncertain prognoses or new treatments, and face-to-face consultations were used instead in such situations ^22)^. Based on these findings, face-to-face explanations are helpful for patient satisfaction when making decisions about very serious, life-altering treatments—even during a pandemic. Conversely, it is not possible to state with certainty that follow-up by video call is not necessary in cases where the illness is not serious. In Japan, telemedicine was promoted as a contingency measure to prevent infection during the rapid spread of COVID-19. Consequently, online and telephonic medical treatment and medication guidance became available to patients who desired it ^23)^. Telemedicine and telenursing have a lower threshold for contact compared to regular telephone consultations and are useful for effective communication and decision-making^24)25)26)^. Additionally, telehealth was an important component of cancer nursing care during COVID-19^27)^, and “provide support via information and communication” was one of the several types of support deemed necessary for discussing patients’ treatment choices during the COVID-19 pandemic. Telehealth is also recommended by the European Society for Medical Oncology^28)^. During a pandemic that requires physical distancing through contact prevention, care using telenursing may support patients’ safe home care. Considering these findings and recommendations, to ensure patient satisfaction with the decision-making process, nurses may be helpful in supporting patients when making a choice between face-to-face or remote care, while reviewing their health status and preferences.

Second, communication skills are important for providing patient-centered care during a pandemic^29)^. One reason why patients in our study continued treatment without being satisfied with their choices was that nurses did not listen to them. This finding suggests that communicative care through listening may reduce the stress caused by patients’ inability to agree on a treatment plan in social situations where changes are needed due to the spread of an infectious disease. Listening is a form of care that attends to the patient’s search for meaning and answers during an illness^29)^. In the Nursing Model for Supporting Shared Decision Making, it is also important to encourage patients to express their feelings and focus on their problems to clarify their true needs and the direction of decision support^30)^. Holistic nursing care that attends to the patient’s suffering also forms the ethical foundation of nursing. To aid patients in making decisions about treatment strategies during a pandemic, it is important to support them in finding meaning in their decisions.

Third, acceptance of recommended treatment was influenced not by patients’ perceptions of their care, but by patients’ feelings of fear of disease progression and anger at not receiving the treatment they desired and their willingness to compromise with the pandemic situation. This is consistent with the results report by Savard et al ^31)^. In their study, patients felt stressed during the COVID-19 pandemic because they believed that changes in cancer treatment schedules and treatments would negatively impact their cancer progression and prognosis. Thus, nurses must support patient decision-making by providing sufficient information to enable the patient to understand their current condition. This includes explaining that the disease may progress if the patient delays seeing a doctor.

Finally, 43.4% of the nurses in our study responded that COVID-19 resulted in a lack of family caregivers’ support. In Japanese medical facilities, patient visits were restricted to prevent the spread of COVID-19. This resulted in a decrease in family caregivers accompanying patients to the hospital to maintain physical distance restrictions and increased burden of care due to caregiver anxiety ^32)^. Family caregiver distress is a dual complication due to both the risk of infection to the patient and also the caregiver’s own social isolation ^33)34)^, and requires the introduction of active nursing support for family caregivers as well as patients. Nursing support for caregivers can be recommended by explaining the use of personal protective equipment, providing remote consultation for caregivers, and establishing social networks on digital platforms^35)36)^.

### Limitations of the study

This study has a limitation in that the survey was not conducted on patients; nurses’ recognition of the patients’ treatment choices was based on nurses’ own assumptions. We propose future prospective observational studies of nurses and patients at the point when a patient needs to change treatment.

### Conclusions

Support of treatment decision-making for patients with cancer during the COVID-19 pandemic included lifestyle advice to prevent infection spread and helping patients understand their choices through listening. By improving the fundamentals of patient-centered care and the ability to respond to infections, these measures may convince patients to accept treatment options recommended by their doctor in future global health crises.

## Data Availability

All relevant data are within the manuscript and its Supporting Information files.

## Acknowledgments

The author would like to thank all the participants in this study.

